# A prospective study of adherence to lenalidomide for multiple myeloma using Medication Event Monitoring System (MEMS) caps

**DOI:** 10.1101/2022.05.30.22275780

**Authors:** Alice E. Silberstein, Mark A. Fiala, Kah Poh Loh, Theresa Cordner, Hira Mian, Tanya M Wildes

## Abstract

**Purpose:** In patients with multiple myeloma, characterizing adherence to orally administered therapies, such as lenalidomide, is critical given their frequent use and potential for poorer outcomes associated with nonadherence. However, little data exist using prospective measures of adherence in this population. Our study piloted use of Medication Event Monitoring System (MEMS) caps and the patient-reported Brief Adherence Rating Scale (BARS) for 3 months in older adults with multiple myeloma.

**Methods:** We enrolled 13 patients with multiple myeloma receiving lenalidomide. Baseline characteristics were summarized; mean adherence to lenalidomide was reported with 95% confidence intervals.

**Results:** The median follow-up was 84 days. Of the 12 participants evaluable, median adherence, as assessed by the MEMS cap data, was 98%. Only 5 had 100% adherence. Deviations from intended use included missed prescribed doses made up during scheduled off week, additional days off between cycles, or taking fewer than anticipated days off. None of these events evident in MEMS data were self-disclosed. The mean difference in adherence estimated between the BARS and MEMS caps was 2%.

**Conclusion:** In this small sample, the observed adherence was higher than reported in retrospective studies using Medication Possession Ratio as a proxy for adherence. The BARS can be easily integrated into clinical encounters but has potential for reporting bias. MEMS caps can help characterize patterns of nonadherence, though there are limitations to their utility and the data can require thorough manual review to reconcile suspected occurrences of nonadherence. Studies should use more than 1 complementary measure of adherence.

Clinicaltrials.gov ID: NCT03779555, Registered 12/19/2018

## Background

Multiple myeloma is the second most common hematologic malignancy and accounts for 1.8% of all new cancer cases [1]. It is a disease of aging, with a median age at diagnosis of 69 years. There has been tremendous progress in therapeutics in multiple myeloma over the past decades with several new drugs being approved. The increasing number of agents and regimens allows for numerous possible regimens and allows providers to tailor treatment based on patient and disease characteristics. Several of the treatments for multiple myeloma can be taken orally, which offer added convenience for patients and potentially higher treatment satisfaction [2]. However, with oral regimens patients must self-manage their medication(s) and nonadherence is a concern.

Lenalidomide is one of the most commonly prescribed treatments for multiple myeloma. Patients often receive lenalidomide during several phases of their myeloma treatment and administration instructions often vary. For example, patients receiving lenalidomide with dexamethasone often take it once daily for 21 days followed by a 7-day break. When co-administered with other treatments, such as bortezomib, it is often administered for 14 days followed by a 7-day break. Following initial treatment, patients may continue on daily maintenance with dose-reduced lenalidomide. If toxicity occurs, a reduction to every other day dosing is not uncommon. The varying of dosing schedules, as well as additional challenges accompanying aging, including comorbidities, polypharmacy, functional limitations, depression, and cognitive changes may impact adherence.

Several retrospective studies have demonstrated that adherence to oral myeloma therapy, and particularly lenalidomide, is a concerning issue, with average adherence rates ranging from 58 to 90% across studies [3-7]. In most prior studies, adherence estimates are based on time between medication refills and the dose supplied in each refill, commonly represented as Medication Possession Ratio (MPR). However, planned treatment pauses, such as the 7-day break common between cycles, and unplanned pauses, such as provider-recommended treatment interruption for toxicity, make MPR an unreliable surrogate for adherence. For example, a patient prescribed 21 days of lenalidomide followed by a 7-day drug holiday would refill a 21-day medication supply every 28 days. Even if that patient were 100% adherent to the prescribed regimen, the maximum MPR for that patient would be 75% (21/28) if the drug holiday is not accounted for. Thus, MPR likely leads to an underestimation of lenalidomide adherence.

Patient-reported adherence has been used as an alternative measure but is also not sufficiently reliable [8, 9]. One recent study in Germany combined patient interviews with prescription data, survey of attending physicians, and caregiver interviews [10]. The researchers found a high rate of adherence. Ninety-seven percent of patients self-evaluated themselves as adherent, defined as always or almost always taking the medication as prescribed. Ninety-eight percent of the patients’ treating oncologists also reported the patient as being adherent. The study demonstrated fairly concordant results with MPR, which had a mean of 99%. However, the validity of patient report and MPR for measuring adherence to lenalidomide is still unclear.

Given these challenges, in the current study we aimed to prospectively examine the rate of adherence of lenalidomide among older adults with multiple myeloma using both objective and subjective approaches to measuring adherence. In addition, we sought to determine if the patient-reported Brief Adherence Rating Scale (BARS) [11] and Medication Event Monitoring System (MEMS) caps would be useful in determining lenalidomide adherence in this pilot study of patients with multiple myeloma in the US.

## Methods

Patients aged 65 years or older receiving lenalidomide-based therapy for multiple myeloma at our institution, a National Cancer Institute (NCI) Comprehensive Cancer Center, were enrolled. Patients were excluded if they met any of the following conditions: had a life expectancy under 6 months; the anticipated duration of lenalidomide therapy was under 3 months; or they did not self-administer their own medications. All participants provided written informed consent. After enrollment participants had a follow-up visit to assess adherence monthly for up to three months. The study was registered with clinicaltrials.gov and approved by the local institutional review board.

Lenalidomide adherence was assessed by MEMS caps, which were affixed to a medication bottle and electronically record every bottle opening. Participants did not have access to the MEMS data. The data from each cap was transferred electronically at scheduled clinic visits and at the end of the study observation period.

Adherence was defined as the proportion of days where adherence was met, either opening the bottle on a treatment day, or not opening the bottle on a nontreatment day. To help identify any inaccuracies in the MEMS data, a self-reported medication diary to record doses skipped, reasons for skipping each dose, and extra cap openings was completed. This diary was turned in at each monthly follow-up assessment. In addition, the participants reported adherence at each follow-up visit using the Brief Adherence Rating Scale (BARS) [11], modified for use with lenalidomide (Supplemental Table). This measure included a visual analog scale to represent the proportion of doses taken in the last month, which was used to quantify patient-reported adherence.

The primary endpoint was mean adherence rate using the MEMS data over the 3-month follow-up period. Assuming a mean adherence of ∼80-100%, *a priori* sample size calculations suggested that a sample size of 45 would allow us to detect mean adherence with a precision of +/- ∼4% with 95% confidence. Secondary analyses included comparisons of the MEMS cap data with the self-reported diaries and patient-reported adherence through descriptive statistics.

## Results

Thirteen participants were enrolled from January to March 2020. At that time, the study was prematurely closed due to institutional policies interrupting research due to the COVID-19 pandemic. Table 1 indicates patient characteristics. The median age at enrollment was 74 years (range 65-92); 9 of 13 were male. Ten participants (77%) identified as non-Hispanic Caucasian and 3 as African-American/Black. Nine participants (69%) were receiving lenalidomide for newly diagnosed multiple myeloma and four (31%) for relapsed disease. Eight participants (62%) were receiving additional antimyeloma treatments in combination with their lenalidomide. At study entry, all participants were on a 28-day treatment cycle where lenalidomide was prescribed for 21 days followed by 7 days off. In addition to antimyeloma treatments, the median number of concomitant medications was 12 (range 2-25).

**Table 1:** Baseline Patient Characteristics.

| Variable Type | N=13 |
| --- | --- |
| Age, years (median, range) | 74 (65-92) |
| *Gender* |  |
| Female | 4 (31%) |
| Male | 9 (69%) |
| *Ethnicity* |  |
| Non-Hispanic Caucasian | 10 (77%) |
| African-American/Black | 3 (23%) |
| *Revised ISS Stage at Diagnosis<sup>1</sup>* |  |
| 1 | 7 (54%) |
| 2 | 4 (31%) |
| 3 | 0 (0%) |
| Unknown | 2 (15%) |
| *Line of therapy* |  |
| First line (induction or maintenance) | 9 (69%) |
| Second Line | 1 (8%) |
| Third Line or Later | 3 (23%) |
| *Lenalidomide dose* |  |
| 2.5 mg | 2 (15%) |
| 5 mg | 3 (23%) |
| 10 mg | 4 (31%) |
| 15 mg | 2 (15%) |
| 25 mg | 2 (15%) |
| *Myeloma Regimens* |  |
| Lenalidomide only | 5 (38%) |
| Lenalidomide, elotuzumab, and dexamethasone | 3 (23%) |
| Lenalidomide, carfilzomib, and dexamethasone | 2 (15%) |
| Lenalidomide, daratumumab, and dexamethasone | 2 (15%) |
| Lenalidomide and ixazomib | 1 (8%) |
| *Number of Comorbidities* |  |
| 0 comorbid conditions | 3 (23%) |
| 1 comorbid conditions | 3 (23%) |
| 2 or more comorbid conditions | 7 (54%) |
| Number of medications (median, range) | 12 (2-25) |
| <sup>1</sup>ISS, *International Staging System* |  |

The median follow-up for adherence was 84 days (range 0-102), including one patient who died due to complications of COVID-19 before any adherence assessments occurred. Of the 12 participants evaluable for adherence, the median adherence assessed by MEMS data was 98% (range 92%-100%; mean = 98%, standard deviation (SD) = 1.9%, 95% CI = 97-99%). Five participants had 100% adherence, including 2 who prematurely discontinued lenalidomide resulting in abbreviated follow-up (26 and 56 days, respectively). In total, 3 patients missed prescribed doses. Of note, 2 of these made up the missed dose during their scheduled off week that cycle. Four patients had at least one additional off day between cycles and 3 skipped at least one scheduled off day. None of the events evident in the MEMS cap data were included on the medication diary or self-reported by the patient to the treating physician or the research staff.

All participants reported strong adherence using the BARS: all reported missing “few, if any (<7) doses” and “never/almost never (0-25% of the time)” taking less than the prescribed number of pills. On the visual analog scale, the average patient-reported adherence was 99.8% (SD 0.3%). Nine of the 13 reported 100% adherence, three reported 99.7% adherence, and one reported 99.0% adherence. Of the 5 participants with 100% adherence assessed by MEMS data, 4 reported 100% on the BARS and 1 reported 99.7%. Of the 7 with <100% adherence assessed by MEMS data, 4 reported 100% adherence on the BARS. The mean discrepancy between MEMS and BARS was 2.0% (SD 2.5%) and the maximum discrepancy was 8%.

Figure 1 depicts the adherence data collected for a participant who experienced multiple changes to the prescribed lenalidomide regimen over the course of the study. During cycle 1, the patient was instructed to take an extra 10 days off (i.e., 17 days off instead of 7) due to an intercurrent acute medical condition. Cycle 2 then proceeded as normal. Beginning in cycle 3, the patient was instructed to switch to every other day dosing during the first 21 days of his cycle, followed by a 7-day holiday as normal. At this point we observed three instances of nonadherence: the patient skipped the last two planned treatments (days 19 and 21 of cycle 3) and then took 8 off days instead of 7. Based on our data, this patient’s adherence rate was 97% because there were 3 days of nonadherence out of 86 days of follow-up. In comparison, estimation of adherence simply based on MPR with no data of prescribed off days would have counted 38 doses out of 86 days followed, yielding an adherence rate of 44%. If prescription data included details of dosing schedules (i.e., the 7-day holiday at the end of the cycle and the switch to every other day dosing), the calculation would have expected 51 doses (35 doses in the first two cycles and 16 in the remainder of the study), and adherence would be estimated at 75%. Even with nuanced data around prescribed dosing schedules, the MPR estimate would be significantly lower because it is unable to account for unplanned holds on medication.

**Figure 1.**
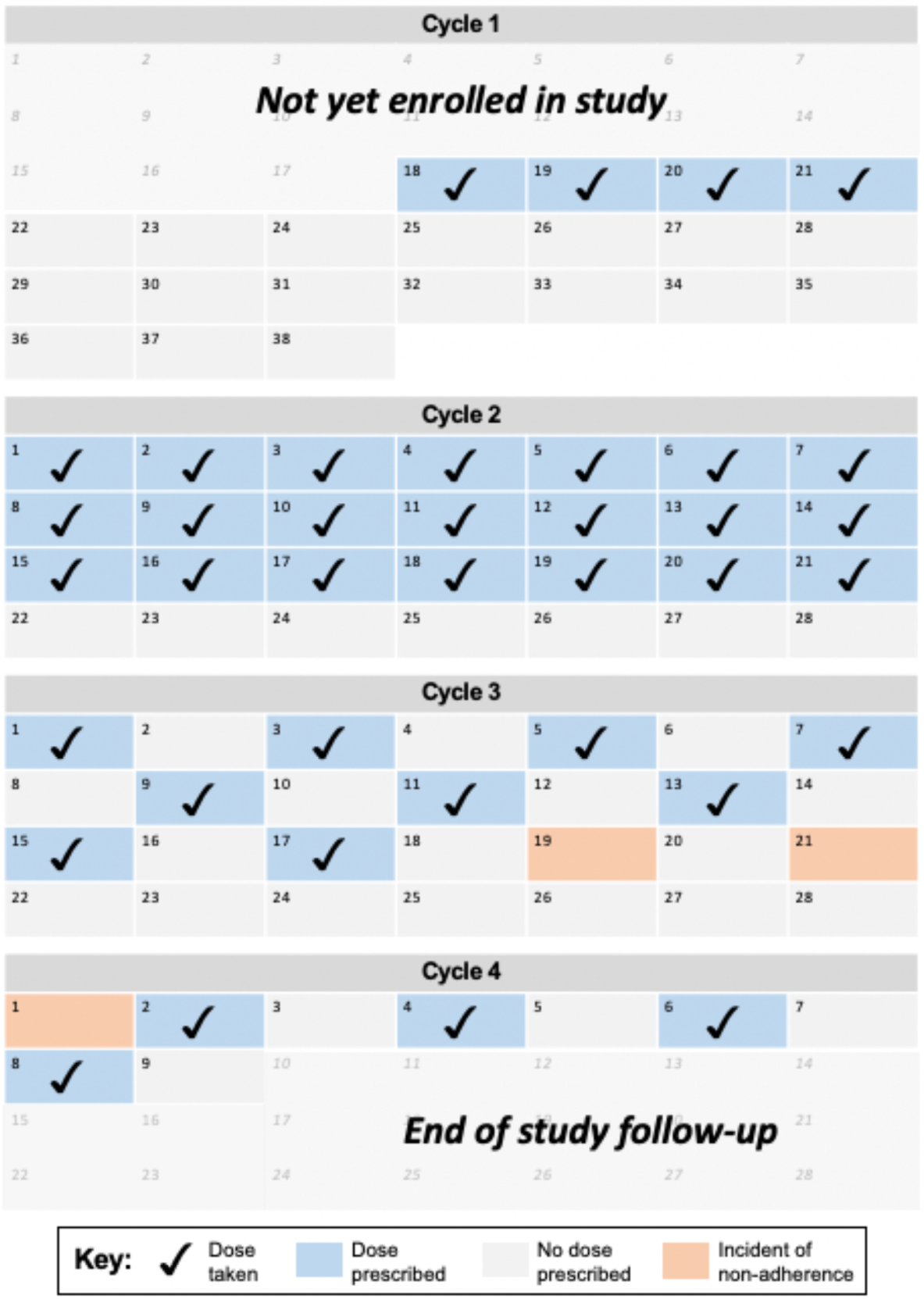
In Cycles 1 and 2 the patient had complete adherence, which included a 10-day treatment break due to toxicity. The dosing schedule was altered in Cycle 3 to every-other-day dosing and the day 19 and 21 planned doses were omitted by the patient. Cycle 4 planned dosing start was delayed by the patient by 1 day for unknown reasons.

While detailed analysis of each MEMS data set yielded a more accurate estimate of adherence compared to MPR, a cursory assessment of the data can result in misinterpretation, particularly when patients take medications at varying times of day. For the most part, the MEMS data could be interpreted simply, where patients were considered adherent either by opening the bottle once on a prescribed treatment day or by not opening the bottle on a prescribed off day. For the study participant represented in Figure 2, the data directly demonstrated adherence in this manner for 88 out of 102 days on the study. However, for the other 14 days there were 7 days where no bottle openings occurred and 7 days where two openings occurred. Examining the raw data demonstrated that the participant typically took each lenalidomide dose late at night, after 10 PM, and occasionally took it later, between 12 and 1 AM. Despite taking medication within the same 2-hour window, this could be interpreted as instances of nonadherence. For our small data set, it was possible to manually adjudicate the data and confirm that this participant had 100% adherence. However, a less detailed analysis would have yielded an adherence rate of 86%, highlighting a potential limitation of using MEMS data on a large scale.

**Figure 2.**
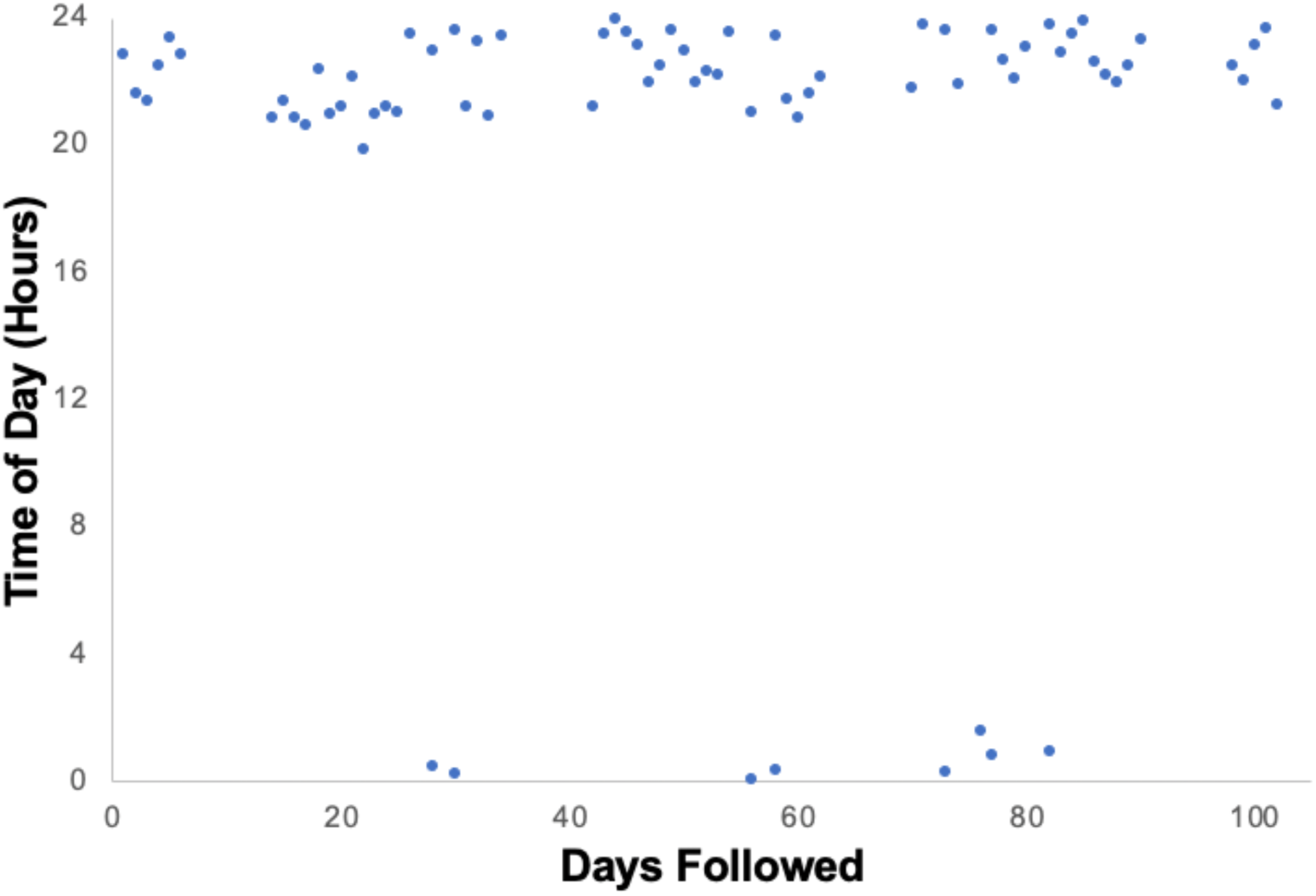
The MEMS data for this patient suggested an adherence of 86% (88/102) as there were 7 planned dosing days where no bottle openings occurred and 7 where two openings occurred. Examining the raw data demonstrated that the patient typically took his pill late at night. In the events that he took it just following midnight, it was reported as nonadherence despite being within an appropriate window for daily dosing.

A third participant’s pattern of adherence data is depicted in Figure 3. While this individual correctly took their lenalidomide on each prescribed day, three instances of nonadherence were observed. For each cycle, the individual only took a 6-day holiday instead of the prescribed 7 days off, beginning the subsequent cycle one day early. The participant was followed for 90 days, yielding an adherence rate of 97%. In comparison, estimating adherence using an MPR approach would have yielded an adherence rate of 100% as there would not be a way to know that the patient started treatment earlier than prescribed. In addition to providing an estimate of adherence rate, the qualitative data from this patient on medication use provides insight into the pattern of adherence. In contrast to the skipped days of treatment seen for the patient in Figure 1, this patient’s nonadherence events had a downstream impact on subsequent cycles, where skipping an “off” day would cause the next cycle to begin and end one day earlier. If such a pattern remained consistent, it could lead to an additional 13 days of treatment per year.

**Figure 3.**
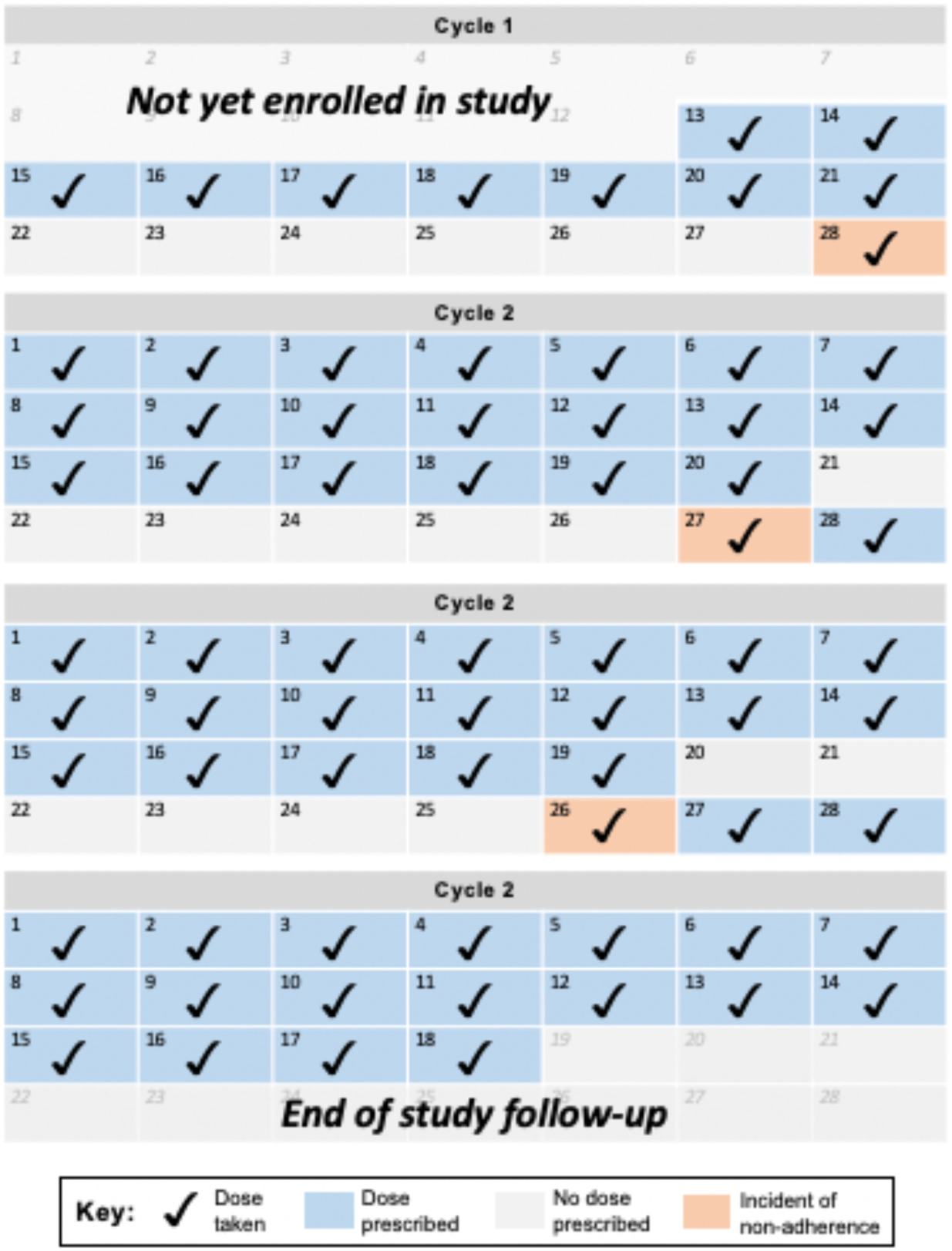
The patient’s lenalidomide was prescribed for 21 days followed by a 7-day treatment holiday. The patient consistently took a 6-day treatment holiday resulting in 3 additional doses being administered over the 90 days of observation.

## Discussion

Our study estimated an average adherence rate of 98% based on MEMS data, which was validated by medication diaries and self-report to the study team. This estimate is higher than that of previous retrospective studies using MPR, but is similar to another prospective study on lenalidomide adherence in myeloma that estimated adherence of 97% [12]. The discrepancy can be partially attributed to the limitations observed with MPR-based approaches. For example, we observed a patient in our study whose adherence by prospective measures was 97%, but whose MPR-based adherence would have been 75% if prescribed drug holidays were accounted for or 44% otherwise, because an MPR approach does not properly account for acute treatment holds. Unplanned and frequent changes may be more relevant among older adults with multiple myeloma who may have increased co-morbidities and high rates of drug toxicity leading to potentially higher rates of treatment regimen and dosing changes.

While MEMS adherence data may more reliably measure adherence compared to MPR in prospective studies, our study also highlights some of the challenges associated with this method. The patient discussed in Figure 2 required manual examination of the data to accurately interpret adherence. This was feasible for our small sample size but would be challenging to scale up and could contribute to occasional measurement inaccuracies, representing a limitation of MEMS data. In addition, we must note that opening a medication bottle or failing to do so is likely an imperfect proxy for taking the medication as prescribed. Lenalidomide is almost uniformly prescribed as one capsule per day, with different strength capsules allowing for different dosages. For medications where more than one capsule/pill/tablet/etc. is required at each dose, MEMS caps would be an insufficient measure for adherence.

Beyond measuring adherence rates in studies, use of MEMS caps may be a way to help patients improve their own adherence. Feiten et al. found that 25% of patients with myeloma found their immunomodulatory drug regimens complicated and that patients with myeloma utilized numerous tools such as pill dispensers, pill package leaflets, and caregivers for help in correctly taking their medications [10]. That study also noted that using a monitoring system may be a method to improve adherence via the Hawthorne effect (alteration of behavior by study subjects due to awareness of being observed) and emphasized the importance of patient understanding of their therapy to support adherence. Therefore, use of the MEMS caps has the potential to improve adherence not only by introducing a monitoring system, but also by supplying a tool that patients could use to keep themselves accountable and that clinicians can use to identify patients who may be misunderstanding their regimen.

In addition to measuring adherence with MEMS data, our study combined use of a diary kept by the patients throughout the month and patient-estimated adherence reported monthly. While the majority of individuals had at least one incident of nonadherence to lenalidomide according to MEMS data, none of the patients disclosed these incidents by either means. The reasons for failing to report nonadherence are unknown: the patients may have been reluctant to admit a mistake, may have misunderstood their dosing schedules, or may have assumed that their nonadherence incidents were minor and not necessary to report. Many patients do not complete the medication diary in real time; rather they complete entries *en masse* later [13]. Because of this, patients may have failed to remember the mistakes.

This study also piloted the BARS, which is a simple assessment that can be integrated into clinical encounters to assess adherence. To date its validity has not been established among a population of people with cancer. Among people with schizophrenia receiving antipsychotic medication, it has been shown to be slightly less accurate than MEMS caps, which is largely considered the gold-standard [11]. In the current study, we found a mean difference of 2% (maximum 8%) in adherence estimates between the BARS and MEMS caps. Unlike the MEMS caps, the BARS self-adherence tool has the advantage of not requiring any additional materials or software to purchase and interpretation does not require any additional review to assess for discrepancies as noted above. However, the BARS is not able to characterize patterns by which nonadherence occurs, cannot assess nonadherence that the patient is unaware of, such as the patient who took one too few off days each cycle noted above, and is prone to bias.

Concerns about adherence are not specific to myeloma; a systematic review of 51 studies of adherence to oral cancer treatments found average adherence rates ranging from 70% to 100% with up to 40% of patients meeting criteria for poor adherence [14]. Given that our study found gaps in reporting of nonadherence in a monitored, highly controlled trial setting, it is likely that similar episodes of nonadherence to oral treatments are occurring across other medical conditions without the awareness of clinicians. These findings suggest that self-report is likely an insufficient method of determining adherence in research or everyday practice. Further, the results call attention to the importance of routine, thorough education of patients around the importance of strict adherence and dosing schedules so that patients are more able to self-identify when instances of nonadherence have occurred.

While in other hematological diseases like chronic myelogenous leukemia the role of nonadherence on poor outcomes is clearly established [16-18], there is a paucity of data regarding the sequelae of nonadherence to treatments for multiple myeloma. One could hypothesize that poor adherence to antimyeloma therapy could be associated with decreased response rates and less durable responses. Gupta et al. examined adherence to oral medications in multiple myeloma in a cross-sectional study and identified associations between lower adherence and lower quality of life as well as greater activity impairment [15]; however, it is unclear if there is any causal link.

In addition to potentially affecting efficacy, poor adherence could also increase the toxicity of lenalidomide. The individual described in Figure 3 of our results provides an example of how adherence could impact outcomes, as the patient was consistently taking six days off from treatment instead of seven. Lenalidomide can cause hematologic toxicity, such as neutropenia and thrombocytopenia [19], and the 7-day drug holiday is designed to limit such toxicities. Taking six days off may not be a sufficient break period and may make the patient more likely to experience toxicity, particularly if this pattern of over-adherence continues over a long period of time.

This study is one of few to prospectively examine adherence in multiple myeloma. We integrated and compared multiple measures of adherence, including MEMS data, patient diaries, and the BARS in patients with multiple myeloma receiving lenalidomide. However, several limitations must be acknowledged. The sample size was small, enrollment was discontinued prematurely due to the pandemic, and follow-up was limited to three months. Thus, adherence rates in the multiple myeloma patient population may differ from those observed in the study. Moreover, prospective adherence studies may be biased to overestimate adherence: voluntary enrollment may be biased toward more adherent patients, or patients may improve adherence by knowing it is being measured [20]. Despite these limitations, this study offers valuable data in a relatively understudied aspect in the care of patients with multiple myeloma.

In conclusion, MEMS data can help assess adherence to oral medications as well as enable one to characterize patterns by which nonadherence occur. Recurring patterns of nonadherence observed within a patient may suggest poor understanding of the prescribed dosing schedule. This stresses the importance of careful education when initially prescribing oral therapies such as lenalidomide and after the prescribed schedule was changed. However, MEMS caps have limitations; the data can require thorough manual review making them potentially difficult to implement on a large scale and they are likely insufficient when multiple capsules/pills/tablets/etc. are required at each dosing. The BARS provides complementary data and can easily be integrated into clinical encounters, but it has the potential for reporting bias, as all self-reported measures do. Additional studies are required to further understand adherence among patients with multiple myeloma and how tools like MEMS caps can be integrated into clinical care, as well as to help identify which patients are at greatest risk for difficulty with adherence and how to best support their adherence.

Clinicaltrials.gov Id: NCT03779555

## Data Availability

All data produced in the present study are available upon reasonable request to the authors

## Acknowledgements

Support: Research reported in this publication was supported in part by the National Cancer Institute of the National Institutes of Health under the Award Number: UG1CA189823 (Alliance for Clinical Trials in Oncology NCORP Grant). https://acknowledgments.alliancefound.org. We would like to thank the Alvin J. Siteman Cancer Center at Washington University School of Medicine and Barnes-Jewish Hospital in St. Louis, Missouri, for the use of the Clinical Trials Core which provided protocol development services. The Siteman Cancer Center is supported in part by an NCI Cancer Center Support Grant #P30 CA91842. The content is solely the responsibility of the authors and does not necessarily represent the official views of the National Institutes of Health.

## Statements and Declarations

### Author Contributions

Mark Fiala, Hira Mian, and Tanya Wildes contributed to the study conception and design. Material preparation and data collection were performed by Mark Fiala and Theresa Cordner, and analysis was performed by Mark Fiala and Alice Silberstein. The first draft of the manuscript was written by Alice Silberstein and all authors commented on previous versions of the manuscript. All authors read and approved the final manuscript.

### Ethics Approval and Informed Consent

The study was registered with clinicaltrials.gov (NCT03779555) and approved by the local IRB. All patients provided informed consent.

---

**Supplemental Table:** Adapted Brief Adherence Rating Scale.

| Item | Response Options |
| --- | --- |
| Over the last month since your last visit, on how many days did you NOT TAKE your lenalidomide? | • Few, if any (<7)<br>• 7-13<br>• 14-20<br>• Most (>20) |
| Over the month since your last visit, how many days did you TAKE LESS THAN the prescribed number of pills of your lenalidomide? | • Always/almost always (76-100% of the time)<br>• Usually (51-75% of the time)<br>• Sometimes (26-50% of the time)<br>• Never/almost never (0-25% of the time) |
| Proportion of doses taken in the last month | Visual analog scale (0%-100%) |

